# The impact of environmental mycobiomes on geographic variation in COVID-19 mortality

**DOI:** 10.1101/2021.12.14.21267549

**Authors:** Joshua Ladau, Katrina Abuabara, Angelica M. Walker, Marcin P. Joachimiak, Ishan Bansal, Yulun Wu, Elijah B. Hoffman, Chaincy Kuo, Nicola Falco, Jared Streich, Mark J. van der Laan, Haruko M. Wainwright, Eoin L. Brodie, Matthias Hess, Daniel Jacobson, James B. Brown

## Abstract

Mortality rates during the COVID-19 pandemic have varied by orders of magnitude across communities in the United States^1^. Individual, socioeconomic, and environmental factors have been linked to health outcomes of COVID-19^2,3,4,5^. It is now widely appreciated that the environmental microbiome, composed of microbial communities associated with soil, water, atmosphere, and the built environment, impacts immune system development and susceptibility to immune-mediated disease^6,7,8^. The human microbiome has been linked to individual COVID-19 disease outcomes^9^, but there are limited data on the influence of the environmental microbiome on geographic variation in COVID-19 across populations^10^. To fill this knowledge gap, we used taxonomic profiles of fungal communities associated with 1,135 homes in 494 counties from across the United States in a machine learning analysis to predict COVID-19 Infection Fatality Ratios (the number of deaths caused by COVID-19 per 1000 SARS-CoV-2 infections^1^; ‘IFR’). Here we show that exposure to increased fungal diversity, and in particular indoor exposure to outdoor fungi, is associated with reduced SARS-CoV-2 IFR. Further, we identify seven fungal genera that are the predominant drivers of this protective signal and may play a role in suppressing COVID-19 mortality. This relationship is strongest in counties where human populations have remained stable over at least the previous decade, consistent with the importance of early-life microbial exposures^11^. We also assessed the explanatory power of 754 other environmental and socioeconomic factors, and found that indoor-outdoor fungal beta-diversity is amongst the strongest predictors of county-level IFR, on par with the most important known COVID-19 risk factors, including age^12^. We anticipate that our study will be a starting point for further integration of environmental mycobiome data with population health information, providing an important missing link in our capacity to identify vulnerable populations. Ultimately, our identification of specific genera predicted to be protective against COVID-19 mortality may point toward novel, proactive therapeutic approaches to infectious disease.

## Introduction

During the first eighteen months of the global COVID-19 pandemic, more than 176 million people were infected with SARS-CoV-2 wich resulted in 3.8 million deaths^13,14^. However, the toll of COVID-19 has varied greatly through both time and across geographical locations: for instance, case-fatality ratios across counties in the United States varied by over four orders of magnitude during the first eight months of the pandemic^15,16,13^. Although some of this variation can be explained by demographic, climatic, or social factors, other factors likely also contribute substantially to area-level variation in COVID-19 disease outcomes^1,17,18^. Identifying these other factors would be of value for forecasting trajectories of this and future pandemics, informing non-pharmaceutical interventions, and potentially indicating research directions towards novel therapeutic and immunological strategies.

In addition to interactions with pathogenic microbes that cause disease, humans interact constantly throughout their lives with a myriad of non-pathogenic microbes (e.g., bacteria and fungi) in the environment^19,20^. These interactions may significantly modulate disease outcomes – for instance, mitigating respiratory illnesses including asthma and allergic disease^21,7,22^. While the mechanisms of action vary, beneficial effects of environmental microbes on health outcomes often share the following characteristics: *(i)* the environmental microbes that have beneficial effects often originate from soils, freshwater environments, plants, and other non-anthropogenic sources, as opposed to the built environment, potentially reflecting a history of human evolutionary adaptation to them^11,7,23,24,25^; *(ii)* environmental microbes reduce disease severity by stimulating immune system development^6,26,11^; and *(iii)* exposure to a diversity of environmental microbes rather than a single taxon is necessary to confer beneficial effects^7,23,25^.

Functional immune response has been reported to be essential for moderating the severity of COVID-19 infection^27,28^. Prompted by the observations that the environmental microbiome influences immune system development and function^6,29^, and that COVID-19 severity correlates with hospital microbiome composition^30^ and with dysbiosis of the lung and^31^ the gut microbiome^32,33^, we hypothesized that the environmental microbiome, specifically the environmental mycobiome, is an important factor determining area-level variation in COVID-19 mortality.

## Results and Discussion

To investigate the link between COVID-19 mortality and environmental fungi, we leveraged data on the taxonomic compositions of fungal communities from 1,135 homes across the United States (Figure E1)^34,35^ and data on COVID-19 from across the United States^13,36^. Each home had paired samples from indoors and outdoors, allowing comparison of the indoor and outdoor fungal communities. Using a novel approach, we also estimated SARS-CoV-2 IFR for each county across the United States, and consequently for each home for which fungal data were available (Supplementary Information). In addition, we extensively explored the influence of demographic, sociological, climate, and soil factors, some of which are known to influence IFR, fungal community composition, or both.

Most people in the United States interact primarily with microbes indoors, spending an average of 87% of their time in their homes and other constructed environments^37^. For fungi, in particular, important differences exist between primarily non-pathogenic outdoor species (which are sometimes also found indoors), and pathogenic or opportunistic species that proliferate in damp indoor environments^38^. Hence, we hypothesized that the presence of outdoor fungi in the built environment may be associated with reduced SARS-CoV-2 IFR. To test this hypothesis, we employed a two-tiered strategy. Firstly, we looked for association between relative fungal abundances (at the genus level) and SARS-CoV-2 IFR using a machine learning approach – iterative Random Forests (iRF). Secondly, we employed a quantile regression strategy to test the prediction that fungal indoor-outdoor beta diversity is most strongly associated with the upper quantiles of IFR – the most severely impacted counties.

The iRF analysis found marginal impact of the relative abundances of fungal genera or indoor-outdoor beta diversity after taking into account 754 demographic, sociological, climate, and soil factors (Supplementary Information). An ablation analysis revealed that after accounting for an expansive collection of other, alternative factors, the impact of fungal genera relative abundances had a statistically significant, albeit small (≈ 1%) effect on mean IFR (Table E1, Figure E7). However, our quantile regression analysis revealed that the 75% and all quantiles above of SARS-CoV-2 IFR were strongly dependent on indoor-outdoor beta diversity. Indeed, indoor-outdoor beta-diversity is associated with reduced SARS-CoV-2 IFR (4.7 or fewer mortalities per 1000 infections; 90th percentile), while dissimilar indoor and outdoor communities (reduced indoor-outdoor beta diversity) are associated with elevated SARS-CoV-2 IFR (12.4 mortalities per 1000 infections; 90th percentile; Figure 1A-B). Hence, while the signal influencing mean IFR is relatively weak, the upper quantiles reveal strong and highly significant associations. These results remained qualitatively the same when the samples from each United States state and Census Division were individually omitted (Figures E2 - E4; Supplementary Information), indicating that the associations we detect are not driven by any single geographic region. Therefore, when accounting for the effects of confounding variables (below), high diversity of outdoor fungi present indoors appears to be associated with suppressed COVID-19 mortality, while lower diversity does not appear to confer these benefits.

**Figure 1:**
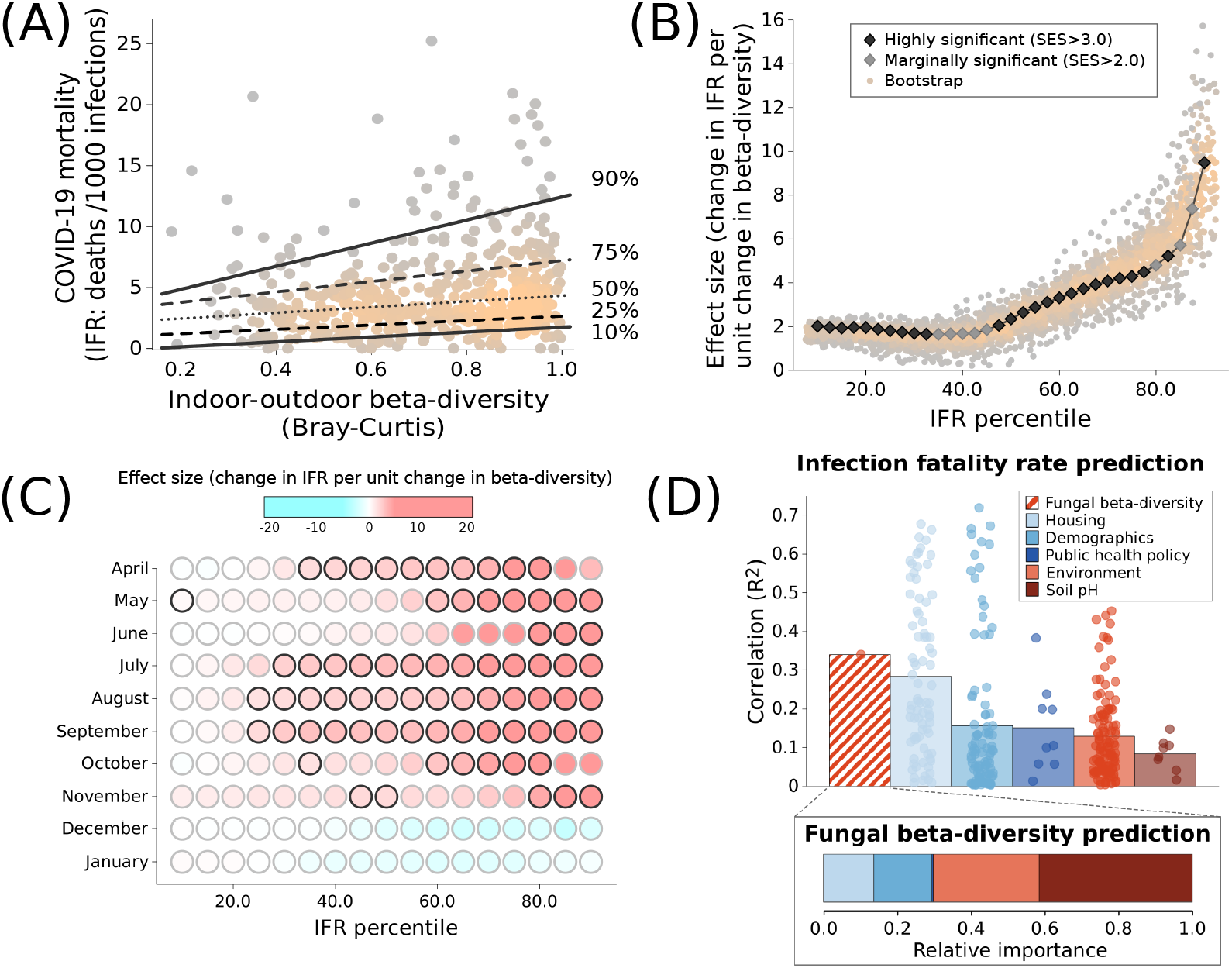
COVID-19 mortality is suppressed in homes where outdoor fungi occur indoors. (A) In United States homes where indoor and outdoor fungal communities are similar (low beta-diversity), COVID-19 mortality is reduced by over a factor of two compared to homes where the communities are dissimilar (high beta-diversity). Each point represents a United States county; shading indicates point density. Although the 90th percentile is sensitive to spatial autocorrelation, these trends are not driven by a single state or region of the United States (Figures E2 - E4). (B) The reductions in COVID-19 mortality (as measured by the standardized effect size, SES) are greatest in the upper quantiles of the COVID-19 mortality distribution, suggesting that outdoor fungi are sufficient but not necessary to reduce COVID-19 mortality. (C) The association between COVID-19 mortality and fungal beta-diversity is strongest early in the pandemic, before December 2020 when vaccination began. Circles with black outlines indicate significant associations (SES> 3). (D) Fungal beta-diversity is a strong predictor of suppression of SARS-CoV-2 IFR relative to other variables [column graph; points represent individual variables, columns show means; correlations give the association between the given variable and the windowed 75th percentile of IFR (see Supplementary Information)]. Moreover, fungal beta-diversity is most strongly associated with soil pH and other environmental variables (inset bar graph), suggesting that it is not a proxy for demographic and other variables that are known to effect COVID-19 mortality.

Multiple lines of evidence indicate that the association between fungal beta-diversity and SARS-CoV-2 IFR suppression may be causal; that is, that the aforementioned associations are driven by the fungi themselves, rather than other confounding factors, such as demographic and climate variables. First, SARS-CoV-2 IFR early in the pandemic (April 2020 - November 2020) is positively associated with fungal indoor-outdoor beta-diversity – when pharmaceutical interventions were unavailable, but the association is weaker later in the pandemic (December 2020 - January 2021) when vaccination began and other pharmaceutical interventions were widely administered (Figure 1C). Second, the differences between indoor and outdoor fungal communities that are associated with SARS-CoV-2 IFR is explained primarily by soil edaphic factors – particularly pH – accounting for over 40% of the variance, while other factors that are known to be related to COVID-19, such as demographic and other non-climate factors, explain less than 30% of the variance combined (Figure 1D). This is intriguing, as it consistent with a causal model where soil edaphic factors affect COVID-19 severity by affecting microbial distributions, which are well-documented to be driven by soil characteristics, including pH^39,40^. Third, causal inference incorporating both fungal indoor-outdoor beta-diversity, demographic, climate, and other predictors indicate a causal link between indoor-outdoor fungal beta-diversity and SARS-CoV-2 IFR even when these other potentially confounding variables are considered (*p* < 0.001, Supplementary Information).

To better understand the components of the fungal communities that contribute to the signal indicating suppression of SARS-CoV-2 IFR we employed a novel, beta-diversity correlation partitioning method (Supplementary Information). At least four of the following fungal genera play a key role in suppressing SARS-CoV-2 IFR: *Alternaria, Aspergillus, Epicoccum, Eurotium, Toxicocladosporium*, and *Wallemia* spp., and a novel Mycosphaerellaceae genus (some of these genera have correlated distributions making effects of individual genera indistinguishable; Figure 2A and 2C). High relative abundance of three of these genera both indoors and outdoors is necessary to detect SARS-CoV-2 IFR suppression: high relative abundance of *Alternaria, Epicoccum*, and Mycosphaerellaceae spp. just indoors – consistent with a primarily indoor origin – or outdoors – consistent with low indoor exposures – is insufficient (Figure 2B and Figure E5).

**Figure 2:**
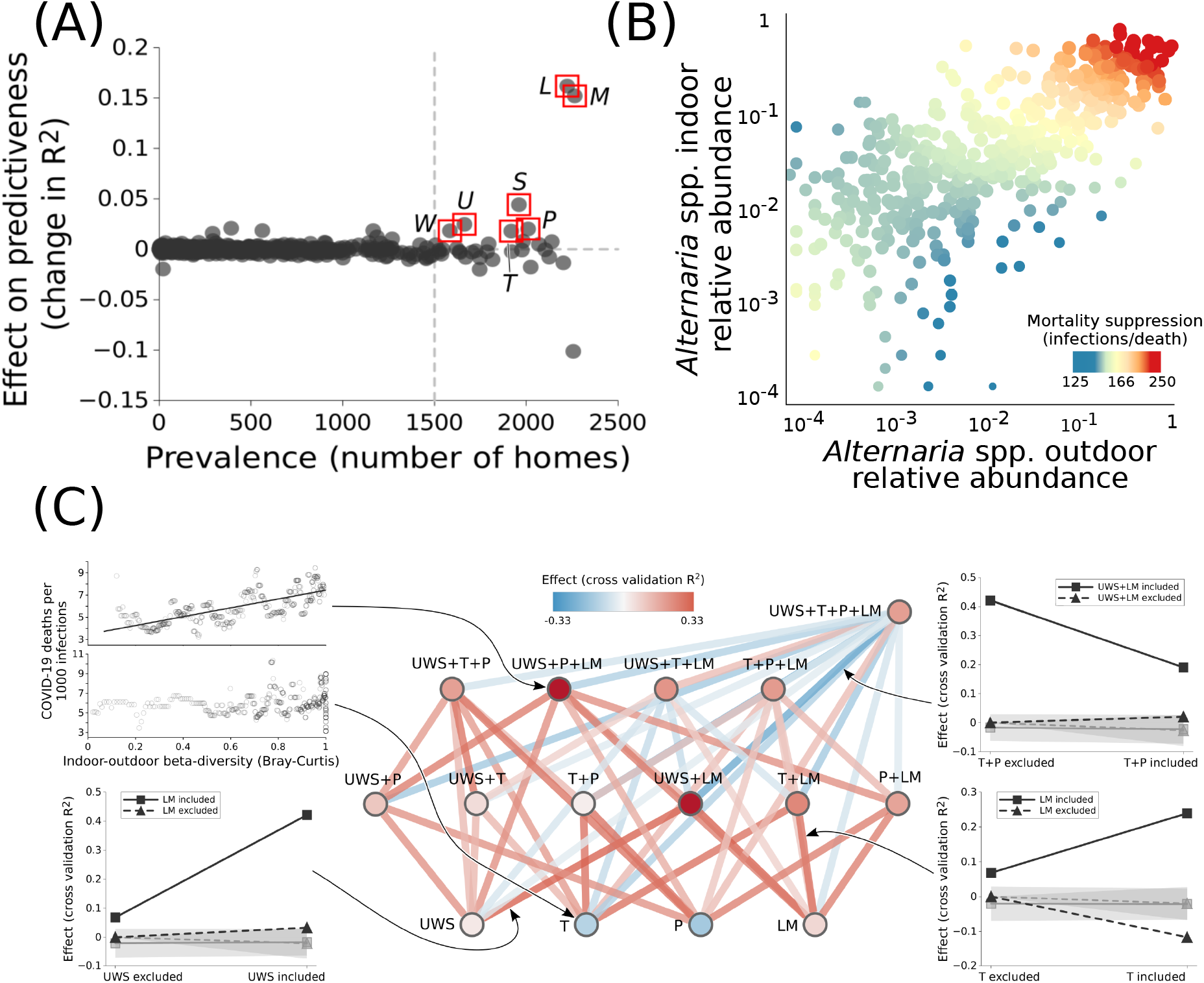
The occurrence of at least four outdoor fungal genera indoors suppresses COVID-19 mortality. (A) Seven prevalent genera significantly increase the predictiveness of indoor-outdoor fungal beta-diversity on COVID-19 mortality. Abbreviations are *L*: *Alternaria, S*: *Aspergillus, P*: *Epicoccum, U*: *Eurotium, T*: *Toxicocladosporium, W*: *Wallemia, M*: Mycosphaerellaceae genus. (B) For some of these genera, including *Alternaria* spp., high relative abundance both outdoors and indoors is necessary for suppressed COVID-19 mortality. (C) However, the full beneficial effects result from synergistic effects of multiple genera: vertices represent sets of genera (colored by predictive power), while edges represent interactions when sets of genera are considered jointly as predictors (colored by interaction strength). Genera with correlated relative abundances are grouped together (e.g., *LM*: *Alternaria* and Mycosphaerellaceae genus). The graphs around the edges show examples of predictive power and interactions; gray lines demarcate null (randomized) expectations plus or minus one standard deviation. Except for when the full set of genera is considered (top right vertex), super-additive effects dominate, pointing to synergistic effects.

Our results pointing to beneficial effects of these seven genera for reducing COVID-19 mortality are novel. Different species from the same genus can have differential effects based on context, for example host genetics and age^41^, health status^42^, and interactions with other microbial exposures^43^. In many contexts, these genera are known to affect health negatively: for example, *Alternaria, Wallemia*, and *Aspergillus* spp. have been associated with increased rates of Irritable Bowel Syndrome, Ulcerative Colitis, and Keratitis, respectively^44,45^ (Table E2). Moreover, two species of *Aspergillus* can worsen outcomes of COVID-19^44^. However, environmental and endemic microbes may also reduce COVID-19 incidence and positively effect COVID-19 outcomes^46,47,48^, and fungi from three aforementioned genera can also improve health outcomes for disease including oral cancer, *Clostridium difficile*-related diseases, and Seborrheic dermatitis, respectively^44^ (Table E2). The mechanism by which fungal elements may reduce infection risk could be through induction of trained immunity^49,29,50^. Fully understanding the context-dependence of the effects of environmental fungi on COVID-19 and other human disease outcomes is an area requiring further study.

In a general context, immune system development has been documented to be promoted by synergistic effects from the exposure to diverse microbes, as opposed to the effects of exposure to individual or a few microbes^7,23,25^. Consistent with these observations, the seven genera that we identified reveal positive synergistic effects: indoor-outdoor differences in relative abundance for genera individually tend to be poor predictors of SARS-CoV-2 IFR suppression (cross validation *R*^2^ between 0 and 0.07, median 0), while the beta-diversity of these genera taken together is strongly predictive (cross validation *R*^2^ between 0.02 and 0.43, median 0.19; Figure 2C).

We hypothesized the association between fungal diversity and SARS-CoV-2 IFR suppression will be strongest in regions where the human population has been stable for the last ten years or more, because people will have been more consistently exposed to the fungi in these regions^6,11,51^. Consistent with this prediction, in locations where the human population remained stable from 2010 to 2017, fungal beta diversity is more strongly associated with SARS-CoV-2 IFR suppression than in regions where it has fluctuated due to population turnover (Figure 3).

**Figure 3:**
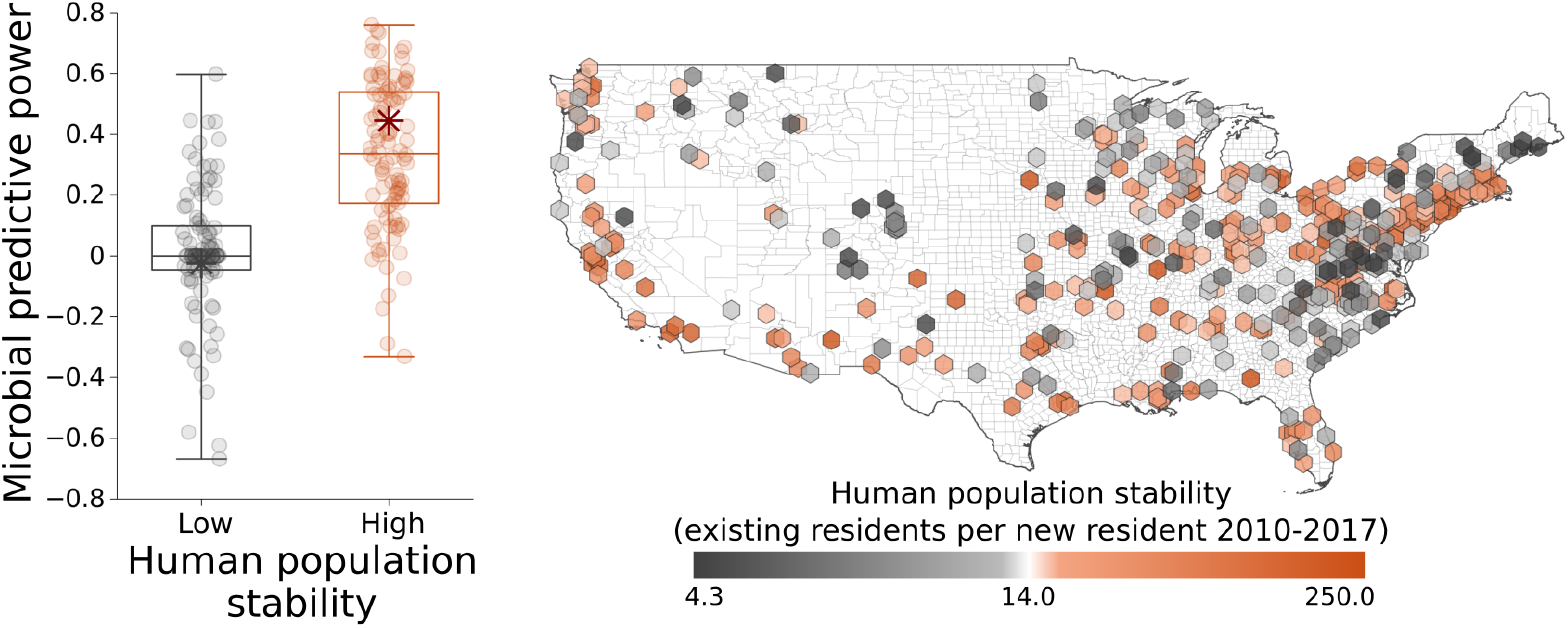
The occurrence of outdoor fungi indoors is more predictive of COVID-19 mortality in locations where people have been less transient from 2010-2017 than in regions where there are many new residents.

Collectively, these results accurately map locations where the occurrence of outdoor fungi in the indoor environment is forecast to most strongly suppress COVID-19 mortality in the United States: the Desert Southwest, Intermountain West, and Upper Midwest; a region that broadly corresponds with part of the country where soils are more alkaline, the primary predictor of indoor-outdoor fungal beta-diversity in our analysis (Figure 4). Soil pH is known to correlate with microbial diversity; it is also related to the water balance of ecosystems (mean annual precipitation relative to mean annual potential evapotranspiration)^52^, which in turn predicts fine dust aerosolization^53^. Thus along with climate^32^, pH may serve as a signature of microbial composition and propensity for microbial transport into homes via dust.

**Figure 4:**
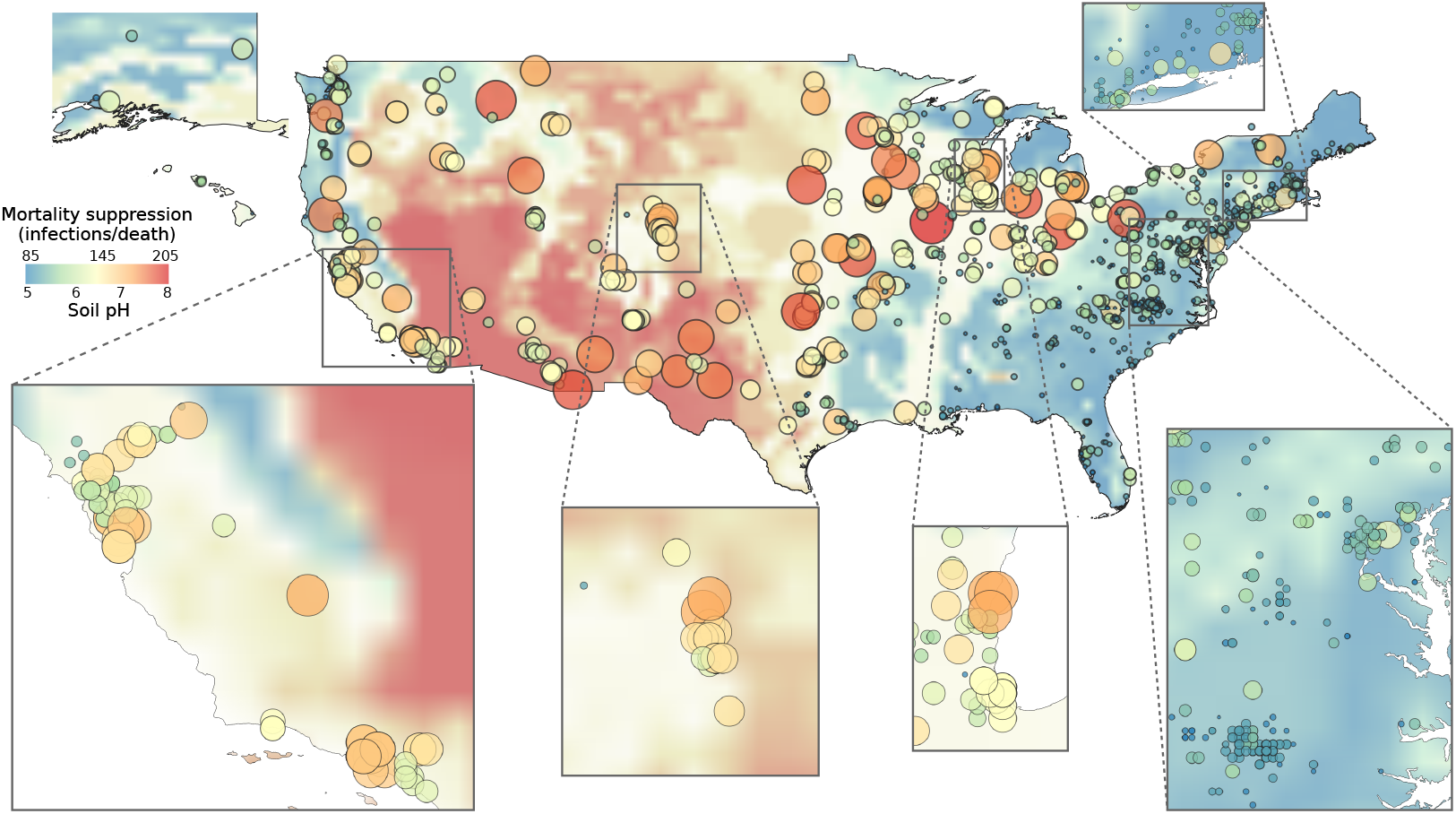
Fungal suppression of COVID-19 mortality varies regionally. In regions where soils tend to be basic (red background shading), indoor-outdoor fungal beta-diversity tends to be low, and fungal suppression of SARS-CoV-2 IFR is high (red dots). By contrast, the opposite trend holds in regions with acidic soils (blue background shading and dots); here, where fungal suppression of SARS-CoV-2 IFR is lessened, SARS-CoV-2 IFR can be high or low depending on whether other factors (e.g., climate, demographics) reduce SARS-CoV-2 IFR.

To reduce fatalities and limit the socioeconomic impact of the COVID-19 and future pandemics, a comprehensive understanding of factors affecting area-level variation in COVID-19 disease outcomes is essential^5^. We develop a novel strategy to address challenges in estimating SARS-CoV-2 IFR (as opposed to the case fatality ratio and other measures of COVID-19 severity), to identify factors driving SARS-CoV-2 IFR at large spatial scales. Here we find that environmental fungal communities, particularly indoor-outdoor beta diversity, are predictive of geographic variation in COVID-19 infection fatality ratios, above and beyond many other social and environmental factors. Our analyses indicate that exposure to high levels of outdoor fungi in homes is protective. If, as supported by our analysis, there is a causal relationship between long-term fungal exposures and SARS-CoV-2 IFR, then the environmental mycobiome constitutes an important missing link in our capacity to identify human populations that are vulnerable to poor outcomes from COVID-19. If, on the other hand, despite our extensive survey of environmental and socioeconomic predictors, we are missing as-yet unidentified confounding factors, our study underscores the utility of the environmental mycobiome as a biosensor^54^. Widespread beneficial effects of environmental fungi may not be specific to COVID-19; limited data support similar findings for childhood allergic disease and other viral diseases^55,56,57^, and it may be relevant for other autoimmune and immune-mediated diseases. Our results provide a foundation for research on the role of fungi and fungal interactions on the immune system, an important addition to a body of literature that has focused primarily on bacteria to date^58,59^. Recent advances in sequencing and classification of environmental fungi will enhance future efforts in this area^60^, and underscore the importance of biosurveillance for this and future pandemics.

## Methods

Details of all methods are provided in Supplementary Information.

## Data Availability

All data produced in the present study are available upon reasonable request to the authors

https://github.com/jladau/Covid19FungiSupplementaryTables

## Acknowledgements

This manuscript has been coauthored by UT-Battelle, LLC under contract no. DE-AC05-00OR22725 with the U.S. Department of Energy. The United States Government retains and the publisher, by accepting the article for publication, acknowledges that the United States Government retains a nonexclusive, paid-up, irrevocable, world-wide license to publish or reproduce the published form of this manuscript, or allow others to do so, for United States Government purposes. The Department of Energy will provide public access to these results of federally sponsored research in accordance with the DOE Public Access Plan (http://energy.gov/downloads/doe-public-access-plan, last accessed September 16, 2020). Work at Oak Ridge and Lawrence Berkeley National Laboratories was supported by the DOE Office of Science through the National Virtual Biotechnology Laboratory, a consortium of DOE national laboratories focused on response to COVID-19, with funding provided by the Coronavirus CARES Act, and was facilitated by previous breakthroughs obtained through the Laboratory Directed Research and Development Program of Lawrence Berkeley National Laboratory. M.P.J. was supported by a grant from the Laboratory Directed Research and Development (LDRD) Program of Lawrence Berkeley National Laboratory under U.S. Department of Energy Contract No. DE-AC02-05CH11231. Chris Mungall and Mark Miller (Lawrence Berkeley National Laboratory) assisted with mapping of Disbiome disease data to the Mondo ontology.

## Author Contributions

J.L., J.B.B., K.A., M.P.J., M.H., Y.W., and I.B. made major contributions to writing the manuscript. J.L., J.B.B., K.A., M.P.J., M.H., Y.W., E.B.H., J.C.R., E.L.B., A.M.C., C.K., A.M.W., N.F., H.M.W., and D.J. designed this study. J.L., J.B.B., K.A., M.P.J., Y.W., E.B.H., I.B. J.C.R., A.M.W., A.M.C., and J.S. contributed analyses. J.B.B., J.L., D.J., and M.L. supervised students and managed analyses.

## Additional Information

Supplementary Information is available for this paper. Correspondence and requests for materials should be addressed to Joshua Ladau.

