## Supplementary Information for "The impact of environmental mycobiomes on geographic variation in COVID-19 mortality"

#### 1 Methods

##### 1.1 Data

###### 1.1.1 Microbial data

Details of sample collection, sequencing, and processing are described in[1,2]. All data are publicly available and were downloaded on 21 July 2020. Briefly, 16S and ITS ribosomal RNA (rRNA) amplicon sequencing were used to profile the bacterial and fungal communities present in dust on the indoor and outdoor door sills of homes across the United States. Because they are typically rarely cleaned, the microbial communities on door sills represent a time-integrated sample of the airborne microbial community at a location. For each home, between 2012-2013, a paired indoor and outdoor sample was collected from each home, allowing a comparison of indoor and outdoor microbial communities. Additional details of sampling collection and processing are Although both bacterial and fungal data were collected, here we focus on the latter; preliminary analyses indicated weaker associations between COVID-19 and bacteria. Each sample was rarefied to 10,000 sequences; samples with fewer than this number of sequences were omitted, yielding paired indoor-outdoor samples from 1,135 homes across the United States.

###### 1.1.2 Raw COVID-19 data

The daily numbers of COVID-19 cases and deaths for each United States county were downloaded from the Johns Hopkins Data COVID-19 data repository[3] on 16 February 2021. The numbers of PCR SARS-CoV-2 tests administered daily in each state in the United States were obtained from the COVID Tracking Project[4] on 16 February 2021.

###### 1.1.3 Estimation of COVID-19 infection fatality ratios

To estimate COVID-19 infection fatality rates in United States counties, two challenges must be overcome. First, because many individuals with SARS-CoV-2 infections are asymptomatic or were not tested, the total number of infections is a latent variable that must be estimated. And second, complicating the latter challenge, the number of tests administered in each

United States county was not systematically recorded, although it was recorded in aggregate across each state. We developed a inferential framework to overcome these challenges, which is detailed in reference[5]. This framework yielded high-quality estimates of the COVID-19 infection fatality ratio ("IFR") in each county for the period beginning 1 March 2020 and ending 31 October 2020. Using this framework, we also calculated IFR estimates for one-month windows beginning on the first day of each month between March 2020 and ending in January 2021.

##### 1.1.4 Demographic and environmental data

To understand the association between of SARS-CoV-2 IFR on fungal beta-diversity, we jointly considered the association of SARS-CoV-2 IFR with 754 other climate, demographic, housing, or COVID-19 policy-related variables ("features") within each United States county. The climate-related features included twelve climate measurements (e.g., average temperature), with monthly breakdowns and 41 static measurements (lacking time-resolution) for a total of 185 features extracted from an extensive literature review[6,7,8,9,10,11,12,13]. The compiled data are described in detail in https://github.com/jladau/Covid19FungiSupplementaryTables. These climate features included global static measures, such as elevation and percent forest cover, as well as seasonal monthly measures, such as maximum temperature and potential evapotranspiration. The 439 demographic-related features included the percentage of individuals in each county for each age group, sex, race, and Hispanic origin; for example, the percentage of Hispanic, white (not in combination with another race) population of females ages 0 to 19 for a given county[14]. The 117 housing-related features captured information relating to the housing units within each county, including data on the structures themselves, as well as the tenants within the homes[15]. The COVID-19 policy-related features included eleven features that captured the percentage of days in the pandemic (between 1 March 2020 and 30 October 2020) that each county had a policy in place, such as a social distancing mandate and the closing of non-essential businesses[16]. This set of features also included two features that capture mask usage, both the chance that a single individual is wearing a mask and an estimate of the number of people wearing a mask in a given county[17]. For a complete list of features see 'feature-descriptions.xlsx' (https://github.com/jladau/Covid19FungiSupplementaryTables).

#### 1.2 Analyses

##### 1.2.1 Overall associations between fungal beta-diversity and COVID-19 mortality

For analyses of diversity, unless specified otherwise, to quantify beta-diversity, the Bray-Curtis dissimilarity of the fungal communities between the inside and outside of each home was calculated. Because the SARS-CoV-2 IFR estimates were at the county grain, dissimilarities from homes within the same counties were averaged to prevent pseudo-replication, non-independence, and under- or over-representation of counties where fewer or more homes were sampled. Overall, this procedure resulted in microbial data for 494 counties across the United States.

Because fungal beta-diversity was expected to be sufficient but not necessary for reducing COVID-19 mortality (i.e., other factors such as demographics are known to reduce mortality), we predicted that the relationship between SARS-CoV-2 IFR and beta-diversity would be "triangular," with consistently low SARS-CoV-2 IFR at low beta-diversity, and both high and low SARS-CoV-2 IFR at high beta-diversity, where reductions may or may not be caused by other factors. Thus, we employed a quantile regression approach, which modeled percentiles of the distribution of SARS-CoV-2 IFR conditional on beta-diversity. We utilized a semi-parametric approach because with our data assumption violations appeared to yield low precision and specificity for standard, normal parametric approaches. Specifically, we first ordered the observations – beta-diversity, SARS-CoV-2 IFR pairs – by beta-diversity. We then calculated the mean beta-diversity and percentile of IFR (10%, 25%, 50%, 75%, 90%) within each possible 20-observation window of the thus ordered data. The relationship between the window means and percentiles was linear, so we fit linear models to measure the association between the percentiles and beta-diversity, and used the slope estimate from these associations as a test statistic. To assess the significance of the slopes, we employed two procedures. First, we employed a randomization test, randomizing the distribution of the raw (not averaged within counties) COVID-19 data, and iterating the entire procedure 100 times to generate a null distribution of slope estimates. Second, we resampled the observations (beta-diversity, SARS-CoV-2 IFR pairs) 100 times with replacement to generate bootstrap distributions of the slope estimates. This quantile regression method was applied in additional analyses described below, using the 75th percentile.

To check the influence of outliers and also investigate the effects of spatial autocorrelation, we recomputed the aforementioned slopes by omitting each state's (fifty iterations) United States Census Division's (nine iterations) samples. If the slopes from these omission analyses were consistent with the slopes from the whole data set analyses, then that would indicate that outliers and spatial autocorrelation were not driving the results observed for the whole data set.

##### 1.2.2 Genera that drive associations between beta-diversity and COVID-19 mortality

To assess which genera drove associations between beta-diversity and SARS-CoV-2 IFR, we used an approach based on hierarchical partitioning. Starting with a single randomly selected genus, we calculated Bray-Curtis dissimilarity. We then calculated the leave-one-out cross validation $R^2$ using the above approach for regressing the 75th percentile of IFR on beta-diversity. With sequential addition of additional genera to the "community" we recalculated beta-diversity, and recorded the corresponding change in cross validation $R^2$ with the addition of each genus. Iterating this process over 100 orderings yielded a mean change in cross validation $R^2$ for each genus. This entire process was also iterated 100 times with the IFR estimates randomized to generate a null distribution of mean change in cross validation $R^2$ statistics for each genus. Genera which (i) on average increased $R^2$ by more than 0.01, (ii) increased $R^2$ significantly ($p < 0.05$) compared to the null distribution of $R^2$ shifts, and (iii) occurred in more than $1,500$ samples (indoor plus outdoor) were identified preliminarily as being drivers of the association between beta-diversity and SARS-CoV-2 IFR. Sets of genera with correlated distributions were clustered using single-linkage clustering on

Spearman-rank correlation coefficients (threshold 0.5). If one or more genus in a cluster was identified preliminarily using the aforementioned procedure, then that *cluster* – i.e., one or more genus in that cluster, but not necessarily the identified genus per se – was inferred to be a driver of the association between beta-diversity and SARS-CoV-2 IFR.

Based on simulations, this approach had low false positive and negative rates at identifying prevalent genera responsible for overall associations between beta-diversity and IFR. To assess error rates, we created 100 simulated data sets where the genera driving the beta-diversity vs IFR association were known. Specifically, we randomly selected half of the genera to be "associated." The beta-diversity of these selected genera was calculated (omitting the other genera) and averaged within each county. To simulate an IFR value for each county, we used a random variate simulated from the following distribution: $0.27 \cdot U \cdot b$, where $U$ is a uniformly distributed random variate on the unit interval, and $b$ is the mean observed beta-diversity over the associated taxa in the county. Iterating this process 100 times generated 100 data sets where the the genera driving the beta-diversity IFR association were "known." For each genus and cluster of genera, we then assessed the confusion matrix for the method outlined above. When clusters, rather than genera per se, were inferred to be associated, the method had low false positive and negative rates (Figure E6A) when genus prevalence was greater than $1,500$ samples. For the genera with prevalence in fewer than $1,500$ samples, despite encompassing over $1,100$ homes, the data set appears to have insufficient power: it appears impossible to reliably identify which of these genera are associated, although false negative rates are controlled for these genera with the implemented method. While this approach toward inferring important genera may seem complex, other, similar and simpler approaches – for instance, when inferences are made about individual genera rather than clusters of genera – have much higher error rates and were prone to false positive results (Figure E6B).

###### 1.2.3 Associations between indoor and outdoor fungal relative abundance, and COVID-19 mortality

To further assess the associations between indoor-outdoor mycobiome and SARS-CoV-2 IFR, we analyzed how IFR was associated jointly with the indoor and outdoor relative abundance of the genera selected using the hierarchical partitioning approach described above. Specifically, we regressed the 75th quantile of IFR on the indoor and outdoor relative abundance of each selected genus (using the R package quantreg[18]). Unlike the analysis of IFR vs beta-diversity, here the assumptions of standard quantile regression methods appeared to be met, and this method also allowed the quantile regression of IFR on two predictors rather than just one, as above. For visualization, we plotted the predicted values of the selected models as a function of the observed indoor and outdoor relative abundances.

###### 1.2.4 Effects of human population transience on associations between fungal beta-diversity and COVID-19 mortality

To assess whether there was an effect of human population transience on the association between fungal beta-diversity and SARS-CoV-2 IFR, we divided the counties into low and high population transience categories based on whether they were in the lower or upper half of the counties ranked by population transience, respectively. Population transience was measured by dividing total immigration to a county over 2014 to 2019 by the 2014 population (data from the American Community Survey https://www.census.gov/programs-surveys/acs/, downloaded on 16 June 2021). The windowed quantile regression analyses described above were performed on each half of the data. In addition, 100 bootstrap iterations were performed to estimate errors.

###### 1.2.5 Correlations between covariates and IFR

For each covariate, we first sort United States counties in descending order relative to the target covariate. Next we applied a sliding window of 20 counties down the rank list, and within each window, we calculated the average value of the covariate, as well as the 75th percentile of IFR. We then fit a linear model and report the $R^2$ values for each pairing.

###### 1.2.6 Machine learning methods for SARS-CoV-2 IFR prediction

Three iterative Random Forest (iRF)[19,20] model configurations were used for this study.

Configuration 1: The first model is configured as follows. Predictors include the following feature sets: a) climate, b) demographic, c) housing, and d) COVID-19 policy features, all at county-level resolution. We use these feature sets as predictors, and fit an iRF model to predict fungal beta-diversity at county resolution. Our goal with this model is to understand which, if any potential confounding factors may contain the same information (e.g., through multi-colinearity) as we have with fungal beta diversity. This is an important analysis, and it complements our causal inference analyses (described below) that also attempts to adjust for all other potentially explanatory features in our dataset.

Configuration 2: The second model configuration predicts SARS-CoV-2 IFR, and uses, as predictors: a) climate, b) demographic, c) housing, d) COVID-19 policy features, e) and fungal beta-diversity along with fungal relative abundances at the phylogenetic level of genera.

Configuration 3: The third model configuration is designed to study the overall feature importance of feature sets a-e above. To do this, we fit iRF models wherein one feature set, e.g., a) climate, was either left-out (omitted), or permuted (each feature within the target feature set is uniformly permuted), and all other features are unperturbed. We then study the effects on predictive accuracy (e.g., median absolute error) after omission or permutation of each feature set. This process is done many times with different random seeds and random permutations to quantify variation—ten sets of five-fold cross validation were generated for a total of fifty model runs per feature set. This process was repeated on each of the five feature sets (a-e, above), and results were compared to the full, unperturbed model.

Down-sampling densely sampled counties: to account for an uneven distribution of fungal samples across United States counties, the model creation included grouped-kfold cross-validation with down sampling. Once for each fold process, the samples in each county were randomly downsampled to five samples. The grouped-kfold sampling process was completed using the GroupKFold function from the sklearn library[21], with an 80/20 split corresponding to five folds. For each model, the five-fold process was repeated ten times, resulting in fifty iRF runs per model. Importantly, results were highly consistent across across runs, with less than 1% standard error in $R^2$ (Coefficient of Determination). For the first fold, sample groups were randomly pulled into the training set until approximately 80% of the samples were included, leaving the other 20% reserved for testing the model. For each subsequent fold, this process was repeated with the caveat of having a completely unique 20% testing set from the previous folds.

Each iRF training run used 1000 trees, five iterations, and 160 threads on the Summit high performance compute system, which is housed at the Oak Ridge Leadership Computing Facility, has approximately 4600 compute nodes, each containing two IBM POWER9 processors, six NVIDIA Tesla V100 GPUs, and 512 gigabytes of RAM. Each iRF testing run used the model created in its corresponding training run, with the 1000 trees and five iterations. The testing run used the fitted iRF models and ran kfold cross-validation to generate beta-diversity and IFR predictions.

##### 1.2.7 Disease associations

To analyze the known microbe-disease interactions of the seven most important fungal taxa, as identified by our hierarchical partitioning analysis, we performed a series of enrichment analyses to evaluate the known associations between these taxa and human diseases for qualitative and quantitative trends. As a quantitative source of correlative disease associations we used the Disbiome resource[22], which collates the associations between 1622 microbes and 372 diseases based on analysis of microbiome data (accessed 13 July 2021). The Disbiome taxa were collapsed to the genus level and each of the 7 fungal taxa (genera) were mapped to any associated diseases in Disbiome. To test for statistical enrichment of specific conditions associated with the seven fungal taxa, we performed a Fisher exact test with a False Discovery Rate (FDR) multiple testing correction on contingency tables based on in and out group counts the seven genera as well as disease associations. In addition to flat taxa-disease associations, we also mapped the Disbiome diseases to the Mondo[23] ontology to further classify and group conditions using their sister and parent relationships. Mapping was performed with Named Entity Recognition using a wrapper wrapped in turbomam/scoped-mapping[24] for the EBI Ontology Lookup Service[25]. With this procedure, certain groups of conditions (such as 'infectious' or by human anatomical site) were tested for statistical enrichment performed as described above. Finally, to identify other possible qualitative trends, additional causally implicated disease associations of the seven fungal species were considered, relying on the DrFungus resource[26]. These were mapped to the Mondo disease ontology, as described above, and were combined into a nonredundant set of taxa to Mondo disease associations based on the union with Mondo disease associations from the Disbiome disease mapping. Frequencies of disease classes were calculated by summing the count of taxa-disease associations per disease category, including multiple disease associations per taxa. Relative disease prevalence was inferred from DrFungus entries as well as clinical reports in the literature. The directionality of correlative effects (positive or negative health effects) was obtained solely from Disbiome data, hence this is also the only source of beneficial effects, and all the other data on fungal associations with diseases was negative[26].

#### 1.3 Code Availability

Except where otherwise noted, all code used to run analyses is available at:

· https://github.com/jladau/COVID19Fungi

· https://github.com/jladau/JavaSource

### 2 Causal Inference: overview of methods and results

To supplement purely correlative analyses, we made use of targeted maximum likelihood estimation (tMLE,[27]), a form of causal inference, to provide an additional line of evidence for detected associations between fungal beta diversity and SARS-CoV-2 IFR. The tMLE framework is particularly useful for studies that make use of machine learning estimators, as tMLE can make use of any predictor (including black box predictors) to inform its estimates of conditional expectations.

The covariate of interest here is fungal beta diversity, which, in causal inference parlance, we call the "exposure variable". Since our exposure variable is continuously valued on [0,1], we design our causal parameter with a stochastic policy to avoid violating positivity of beta diversity scores. Essentially, we assess the causal effect based on the counterfactual outcomes under exposure levels drawn from counterfactual distributions shifted from the original distribution of the exposure variable.

The intuition is, if we observe notable differences between average outcomes under more aggressive/conservative exposure assigning policies, then we have more confidence to draw the conclusion that the exposure has a significant causal influence on the response. This framework is known as stochastic intervention and its usage in inference has been well established in previous studies[28,29]. For the purposes of model interpretation, we designed a summary parameter via a working Marginal Structural Model[30] to statistically characterize the positive linear trend observed in this figure, and presented the results in section 2.1. Note that the goal of designing this parameter is to make statistical inference and provide interpretability — we do not assume a linear diversity-IFR relation.

We used a doubly robust estimation framework – Targeted Maximum Likelihood (TML)[27] to estimate the counterfactual expected outcomes and construct their asymptotic normal distributions. Then, we derived the estimation and confidence interval of the summary linear coefficient between IFR and beta diversity via the Delta method. The TML estimators allow more relaxed convergence conditions compared to plug-in and inverse probability weighted (IPTW) estimators, and are asymptotically efficient[27]. The standard deviations of the estimators can be analytically derived from the efficient influence curve[31] which allows us to construct confidence bonds. The underlying conditional density of beta diversity and conditional expectation of IFR are fitted with Super Learning[32], which builds an ensemble model as a weighted combination of predictive functions from a diverse collection of statistical and machine learning algorithms.

#### 2.1 Evidence that the associations between microbial beta-diversity and COVID-19 severity may be causal

Finally, causal inference incorporating a) climate, b) demographic, c) housing, d) COVID-19 policy features, e) and fungal beta-diversity along with fungal relative abundances at the phylogenetic level of genera point to a strong causal link between indoor-outdoor microbial beta-diversity and SARS-CoV-2 IFR even when these other potentially confounding variables are considered. The estimations of expected 75-quantile IFR under counterfactual beta diversity distributions are shown in Figure E7, and the table below shows the estimations of the linear coefficient presented in this figure. The last column is the confidence level to reject the null hypothesis that there is no positive linear trend in 75-quantile IFR with an increasing fungal beta diversity score—which can be thought of as similar in purpose to a $p$-value.

| | Plug-In Estimator | TML Estimator | TMLE Std | Confidence level (1 − p-value) |
|---|---|---|---|---|
| Mar.2020-Oct.2020 | $7.43e-04$ | $6.68e-04$ | $3.72e-05$ | 100.00% |
| Mar.2020-Feb.2021 | $1.67e-04$ | $9.98e-05$ | $3.75e-05$ | 99.61% |
| Mar.2020-Oct.2020 w/ selected fungi | $5.10e-04$ | $5.02e-04$ | $4.97e-05$ | 100.00% |
| Mar.2020-Feb.2020 w/ selected fungi | $1.06e-04$ | $9.82e-05$ | $5.09e-05$ | 97.32% |

The results confirm that there is a significant positive increment in the outcome when the exposure distribution is shifted rightwards, meaning that higher beta diversity scores may *cause* higher 75-quantile IFR on the population. Please note, we take this only as another line of evidence. While the rapidly growing field of causal inference is compelling, it is no substitute for counterfactual experiments. However, when counterfactual experiments are difficult or impossible, as may be the case here, causal inference procedures, including tMLE, are useful to supplement purely correlative analyses.

#### 2.2 Causal inference: background and methods

In a large number of causal inference problems, researchers design a causal parameter whose estimation is based on some sort of static counterfactual mechanism, meaning that the parameter relies on the distribution of outcome $Y$ given that the exposure variable $A$ is deterministically set to some specific alternative values. A classic example is the estimation of average treatment effect (ATE) using a directed acyclic graph (DAG) model in a clinical trial with binary intervention.

In such model settings, one often has to fit the data generating mechanism with suitable statistical learning methods and assume a given individual's likelihood of having an alternative exposure level $a$ (within the range of consideration) is always positive. Although this assumption is reasonable in the example above, it becomes highly unrealistic in a setting where the exposure variable is highly diverse or even continuous.

One way to address this issue, presented by Díaz and van der Laan[28], is to consider a set of counterfactual exposure assigning policies under shifted distributions. This technique is referred to as stochastic interventions. We use this approach to model the population causal effect of fungi's beta diversity level on COVID-19's fatality rate.

Since the 75-quantile IFR can not be directly observed, we take the route of fitting a quantile model with a portion of our data and acquiring the 75-quantile IFR for the subjects in the remaining portion. This remaining portion of the data is used for causal inference.

#### 2.3 Causal Inference: Parameters of Interest – Stochastic Interventions

Let $O = (W, A, Y)$ be the observation variable consisting of the confounding, exposure and outcome triple, $P = (p(W), p(A \mid W), p(Y \mid A, W))$ be the true probability model. $E_{A \sim p_\delta}(Y)$ denotes the average outcome under an exposure distribution $p_\delta(A \mid W)$, then we let $\{p_\delta(A \mid W) = p(A - \delta(A, W) \mid W)\}_\delta$ be a family of density functions shifted from the true density $p$, in which $\delta(A, W)$ is given by

$$\delta(a, w) = \begin{cases} \delta & \text{if } a > l(w) + \delta \text{ and } a < u(w) + \delta \\ a - l(w) & \text{if } a \leq l(w) + \delta \\ a - u(w) & \text{if } a \geq u(w) + \delta \end{cases} \tag{1}$$

where $[l(w), u(w)]$ is the range of $A$ given $w$. To clarify, each $p_\delta(A \mid W)$ within the family is a counterfactual distribution constructed via right-shifting the original distribution by $\delta$, with a cap on the shift so that its support does not go beyond the permissible range of $A$. In a nonparametric structural equation model (NPSEM,[33]) setting, the corresponding family of parameters $\{\Psi(\delta) = E_{A \sim p_\delta}(Y)\}_\delta$ can be identified under standard randomized assumption $A \perp Y_a \mid W$ by

$$\Psi(\delta) = \int_{\mathcal{W}} \int_{\mathcal{A}} E(Y \mid a, w) p_\delta(a \mid w) p(w) \, d\lambda(a) \, d\mu(w) \tag{2}$$

$$= E\left[E\left[E(Y \mid A + \delta(A, W), W) \mid W\right]\right] \tag{3}$$

where $\lambda$ and $\mu$ are suitable measures for $a$ and $w$ respectively. These parameters reflect the variation of resulting outcomes given different levels of exposure intensity, and the robust estimation of these parameters involves propensity weights $p(A - \delta(A, W) \mid W)/p(A \mid W)$ in which the discrepancy between the numerator and denominator can be decently smaller than the ones under deterministic policies. To statistically characterize this influence of $\delta$ over $Y$, we design a summary parameter via a working Marginal Structural Model (MSM,[30]) as follow

$$\beta^* = \arg\min_\beta \int_{\mathcal{D}} \left[\Psi(\delta) - (\alpha + \beta \cdot \delta)\right]^2 dv(\delta) \tag{4}$$

where $v$ is a suitable measure, e.g. a discrete uniform over a grid of customized $\delta$ values. This parameter captures the positive/negative linear trend in the estimated mean outcomes under shifting exposures.

#### 2.4 Causal Inference: Estimations – Targeted Maximum Likelihood

In evaluating $\Psi(\delta)$, we apply the Targeted Learning framework[27]. Under some regularity assumptions[28], the tMLE estimator $\hat{\Psi}_n(\delta)$ will be consistent as long as either one of $E(Y \mid A, W)$ and $p(A \mid W)$ is estimated consistently. The estimator is asymptotically normal if the model fits $\hat{E}_n(Y \mid A, W)$ and $\hat{p}_n(A \mid W)$ have a combine order of L-1 error less than $n^{-0.5}$ (e.g. $n^{-0.25}$ each) as compared to the plug-in or inverse probability weighted (IPTW) estimator for which $\hat{E}_n(Y \mid A, W)$ or $\hat{p}_n(A \mid W)$ must have an order of error $< n^{-0.5}$ individually. Figure E8 provides a demonstration of the need for TMLE. Once these assumptions hold, the estimator is asymptotically efficient, and the variance can be derived and estimated by its efficient influence curve (EIC) denoted as $D_P(O)$:

$$D_P(O) = \frac{p(A - \delta(A, W) \mid W)}{p(A \mid W)}[Y - E(Y \mid A, W)] + E(Y \mid A + \delta(A, W), W) - \Psi(\delta) \quad (5)$$

with which the asymptotic distribution of $\hat{\Psi}_n(\delta)$ can thus be constructed as

$$\sqrt{n}(\hat{\Psi}_n(\delta) - \Psi(\delta)) \Rightarrow \mathcal{N}(0, \Sigma) \quad (6)$$

where $\Sigma = E\left[D_P(O) \cdot D_P(O)^T\right]$ and can be estimated by

$$\hat{\Sigma}_n = E_n\left[D_{\hat{P}_n}(O) \cdot D_{\hat{P}_n}(O)^T\right] = \frac{1}{n}\sum_{i=1}^{n}\left[D_{\hat{P}_n}(O_i)\right]^2 \quad (7)$$

Upon attaining the estimates of $\Psi(\delta)$, the asymptotic distribution of the slope estimate $\hat{\beta}_n^*$ is derived with the Delta method, with which we evaluate the $p$-value of the null hypothesis—that there is no positive linear trend in 75-quantile IFR given a shifting fungal beta diversity score. The confidence level of rejecting this null hypothesis thus follows.

### 3 Results

#### 3.1 Disease Associations

Beneficial environmental microbiome effects on human health are less frequently reported than disease, however the Disbiome[22] resource is the first collection of correlative human microbiome taxa associations with human diseases. While the human and environmental microbiomes are distinct measurements, there is data that environmental microbes are reflected in human microbiome taxa abundance[34,35]. We observed that the seven fungal genera can originate from a variety of indoor and outdoor habitats, including soil, plants, lakes, and food spoilage[26]. For these genera we compiled taxa-disease associations across Disbiome data and curated data on mycoses[26] and also categorized them into disease classes by their Mondo ontology parent terms (https://github.com/jladau/Covid19FungiSupplementaryTables). Many of these genera and species can be associated with numerous effects on human diseases, beneficial or harmful, including *Clostridium difficile*-related diseases, Caries, and Crohn's diseases[22 26] (Table E2). Our knowledge of causal fungal-disease interactions is still incomplete but based on existing microbiome and case study data we wanted to check whether any of these genera are known to affect human health. Half of the disease associations are with the inflammatory, digestive, and respiratory disease classes, often for conditions that are considered allergic reactions or autoimmune responses. 20% of the associations are with inflammatory or autoimmune diseases (see Methods), suggesting a direct link to aspects of the immune system not necessarily involved in response to infections, although infectious disease associations also appear to be enriched ($p$=0.025; Table E3), however our enrichment analysis does not include the direction of the effect of taxa presence on disease outcome. Since our knowledge of human diseased, environmental microbes, and their interaction are incomplete, it is possible that the associated non-infectious diseases have an infectious component, or that there are unrecognized aspects of immune response or health status in what are currently considered infectious diseases. Some of the respiratory or other allergies related to these fungal genera could be under-reported if symptoms are mild, transient, or misdiagnosed. These disease class associations results show that fungal diseases often occur in tissues exposed to the environment (digestive, respiratory, intestinal), and are not involved with cardiac, renal, or hematologic systems. Several diseases were associated with multiple of the seven taxa, most notably *Clostridium difficile* associated disease, Caries, and Crohn's disease all having associations to three of the seven taxa. Interestingly, these three conditions occur in environmentally exposed human tissues, and specifically the gastrointestinal system, which is consistent with environmental microbe exposure.

We considered one more level of information for Disbiome taxa to disease associations, and that is the direction of the effect on disease out comes. In many cases Disbiome provides an 'elevated' or 'reduced', and we find many positive interactions between the seven genera or species belonging to that genera and human diseases (Table E2). Overall, while the total number of negative fungal associations outnumbered positive ones, we believe this is partly due to the biased focus on negative disease outcome in current metagenome and clinical studies, which in general do not focus on salutology. Finally, we observe that *Aspergillus* species are associated with both positive and negative disease outcomes, involving different diseases and different *Aspergillus* species. Hence, members of the same genus can have both positive and negative effects depending on the disease condition, and likely other factors such as human genotype[36]. Two *Aspergillus* species were associated with worse COVID-19 outcomes, however we believe this is influenced by bias from negative studies not allowing to observe more positive associations and also that early protective exposure to fungi is likely to be independent from presence of these fungi in the human microbiome later in life (e.g., Disbiome data). Thus we are still limited in our knowledge of fungal disease associations, with 28 disease associations for the seven fungal genera associated with lower IFR, given that these genera correspond to thousands or more species. However, we do know that fungi are abundant in indoor and outdoor environments and rarely cause serious disease but are common opportunistic pathogens. Given this, our ability to derive specific and causal hypothesis for fungal affects on human disease remains limited warranting further human fungal sequencing studies.

[6] Gunther Fischer, Freddy O Nachtergaele, Sylvia Prieler, Edmar Teixeira, Geza Toth, Harry van Velthuizen, Luc Verelst, and David Wiberg. *Global Agro-ecological Zones (GAEZ v3.0) Model Documentation*. International Institute for Applied Systems Analysis (IIASA) and the Food and Agriculture Organization of the United Nations (FAO), Laxenburg, Austria and Rome, Italy, 2012.

[7] Robert J Zomer, Antonio Trabucco, Deborah A Bossio, and Louis V Verchot. Climate change mitigation: A spatial analysis of global land suitability for clean development mechanism afforestation and reforestation. *Agriculture, ecosystems & environment*, 126 (1-2):67–80, 2008.

[8] Robert J Zomer, Deborah A Bossio, Antonio Trabucco, Li Yuanjie, Diwan C Gupta, and Virendra P Singh. *Trees and water: smallholder agroforestry on irrigated lands in Northern India*, volume 122. IWMI, 2007.

[9] Guenther Fischer, Freddy Nachtergaele, Sylvia Prieler, HT Van Velthuizen, Luc Verelst, and David Wiberg. Global agro-ecological zones assessment for agriculture (gaez 2008). *IIASA, Laxenburg, Austria and FAO, Rome, Italy*, 10, 2008.

[10] IGBP-DIS. Soildata (v. 0). a program for creating global soil-property databases. *IGBP Global Soils Data Task*, 1998.

[11] Stephen E Fick and Robert J Hijmans. Worldclim 2: new 1-km spatial resolution climate surfaces for global land areas. *International journal of climatology*, 37(12):4302–4315, 2017.

[12] Adam M Wilson and Walter Jetz. Remotely sensed high-resolution global cloud dynamics for predicting ecosystem and biodiversity distributions. *PLoS biology*, 14(3): e1002415, 2016.

[13] A Trabucco and RJ Zomer. High-resolution global soil-water balance explicit for climate–standard vegetation and soil conditions. *CGIAR Consortium for Spatial Information*, 2010.

[14] United States Census Bureau. *Annual County Resident Population Estimates by Age, Sex,Race, and Hispanic Origin: April 1, 2010 to July 1, 2018 (CC-EST2018-ALLDATA)*, 2020. URL https://www.census.gov/data/tables/time-series/demo/popest/2010s-counties-detail.html. Accessed: June 1 2020.

[15] United States Census Bureau. *2018 American Community Survey 5-Year Estimates Data Profiles*, 2020. URL https://data.census.gov/cedsci/map?q=Housing%20Units%20in%20the%20United%20Statesg=0 Accessed: June 1 2020.

[16] Keystone Strategy. *Keystone-Strategy/covid19-intervention-data*, 2020. URL https://github.com/Keystone-Strategy/covid19-intervention-data. Accessed: October 30 2020.

[17] The New York Times. *nytimes/covid-19-data/mask-use*, 2020. URL https://github.com/nytimes/covid-19-data/tree/master/mask-use. Accessed: July 28 2020.

[18] Roger Koenker, Stephen Portnoy, Pin Tian Ng, Achim Zeileis, Philip Grosjean, and Brian D Ripley. Quantile regression. URL https://www.r-project.org/.

[29] Nima S Hejazi, Mark J van der Laan, Holly E Janes, Peter B Gilbert, and David C Benkeser. Efficient nonparametric inference on the effects of stochastic interventions under two-phase sampling, with applications to vaccine efficacy trials. *Biometrics*, 2020.

[30] James M Robins, Miguel Angel Hernan, and Babette Brumback. Marginal structural models and causal inference in epidemiology, 2000.

[31] Peter J Bickel, CAJ Klaassen, Y Ritov, and JA Wellner. Efficient and adaptive inference in semiparametric models, 1993.
