## Extended Data Figures and Tables for "The impact of environmental mycobiomes on geographic variation in COVID-19 mortality"

| Model | (Mean AE)/Mean | MSE | $R^2$ | (Median AE)/Median |
|---|---|---|---|---|
| Fun + Dem + Cli + Hou + Pol | 0.39739 | 7.1159e-06 | 0.4141 | 0.3378 |
| Dem + Cli + Hou + Pol | 0.4154 | 7.5844e-06 | 0.4062 | 0.3707 |
| Fun + Cli + Hou + Pol | 0.4015 | 7.0784e-06 | 0.4085 | 0.3432 |
| Fun + Dem + Hou + Pol | 0.4089 | 7.2851e-06 | 0.3946 | 0.3405 |
| Fun + Dem + Cli + Pol | 0.3989 | 6.6547e-06 | 0.4239 | 0.3306 |
| Fun + Dem + Cli + Hou | 0.4061 | 7.7363e-06 | 0.3932 | 0.3437 |
| Fun (shuffled) + Dem + Cli + Hou + Pol | 0.39737 | 7.1208e-06 | 0.4139 | 0.3313 |

Table E1: Feature-class ablation analysis. Each feature-class, Fun (fungal beta-diversity), Dem (demographic), Cli (climate), Hou (housing), Pol (COVID-19 policy), is ablated one at a time, and measures of accuracy and variability are reported. In addition to ablation analysis, we study the effects of feature permutation on Fungal feature (Fun), and find that the permuted features out-perform (on average), un-permuted features, indicating that while including Fungal (Fun) features in the model improves predictive performance, this improvement is not dependent on the joint distribution of Fungal features, only on the marginal distribution. AE and MSE stand for absolute error and mean standard error, respectively.

| Genus | Disease Name | Organism Name in Disbiome | Disease Outcome |
|---|---|---|---|
| *Alternaria* | Crohn's Disease | *A. brassicicola* | Elevated |
| *Alternaria* | Irritable Bowel Syndrome | *A. alternata* | Elevated |
| *Alternaria* | Oral lichen planus | *Alternaria* sp. | Elevated |
| *Alternaria* | Vogt-Koyagani-Harada disease | *Alternaria alternata* | Elevated |
| *Wallemia* | Ulcerative Colitis | *Wallemia* sp. | Elevated |
| *Aspergillus* | Crohn's Disease | *A. clavatus* | Elevated |
| *Aspergillus* | Dandruff | *Aspergillus* sp. | Elevated |
| *Aspergillus* | Keratitis | *Aspergillus* sp. | Elevated |
| *Aspergillus* | Psoriasis | *Aspergillus* sp. | Elevated |
| *Aspergillus* | Uveitits | *A. gracilis* | Elevated |
| *Aspergillus* | Pediculosis | *A. fumigatus* | Elevated |
| *Aspergillus* | Oral lichen planus | *Aspergillus* sp. | Elevated |
| *Aspergillus* | Cystic fibrosis exacerbation | *Aspergillus* sp. | Elevated |
| *Aspergillus* | Covid-19 | *A. flavus* | Elevated |
| *Aspergillus* | Covid-19 | *A. niger* | Elevated |
| *Alternaria* | HIV infection | *Alternaria* sp. | Reduced |
| *Alternaria* | Oral cancer | *A. tamaricis* | Reduced |
| *Alternaria* | Oral cancer | *A. alternata* | Reduced |
| *Alternaria* | Caries | *A. alternata* | Reduced |
| *Wallemia* | *C. difficile* associated disease | *W. mellicola* | Reduced |
| *Wallemia* | Crohn's Disease | *Wallemia* sp. | Reduced |
| *Aspergillus* | *C. difficile* associated disease | *A. penicillioides* | Reduced |
| *Aspergillus* | *C. difficile* associated disease | *A. austroafricanus* | Reduced |
| *Aspergillus* | Psoriasis | *Aspergillus* sp. | Reduced |
| *Aspergillus* | Seborrheic dermatitis | *Aspergillus* sp. | Reduced |
| *Aspergillus* | Caries | *A. sydowii* | Reduced |
| *Epicoccum* | Alcoholism | *Epicoccum* sp | Reduced |
| *Toxicocladosporium* | Caries | *T. strelitziae* | Reduced |

Table E2: Microbiome taxa correlated with diseases along with direction of the effect, for all seven fungal genera implicated in reducing COVID-19 mortality. Both genera and species-level associations are shown. See Supplementary Information for references.

| **Rank** | **Disease Class** | **Frequency** | **P-Value** | **Associated Genera** |
|---|---|---|---|---|
| 1 | Infectious | 0.19 | 0.02512 | Mycosphaerellaceae spp., *Alternaria*, *Wallemia*, *Aspergillus* |
| 2 | Inflammatory | 0.17 | 0.6879 | Mycosphaerellaceae spp., *Alternaria*, *Eurotium*, *Wallemia*, *Aspergillus* |
| 3 | Digestive | 0.17 | 1 | Mycosphaerellaceae spp., *Alternaria*, *Wallemia*, *Aspergillus*, *Toxicocladosporium* |
| 4 | Respiratory | 0.16 | 0.7056 | *Alternaria*, *Eurotium*, *Wallemia*, *Aspergillus* |
| 5 | Intestinal | 0.05 | 0.6464 | *Alternaria*, *Wallemia*, *Aspergillus* |
| 6 | Autoimmune | 0.03 | 1 | Mycosphaerellaceae spp., *Alternaria* |
| 7 | Endocrine | 0.02 | 1 | *Aspergillus* |

Table E3: Disease associations for the seven fungal genera implicated in reducing COVID-19 mortality. Frequencies of the association for each Mondo ontology disease class are shown along with statistical significance of enrichment based on contingency table analyses. See Supplementary Information for details.

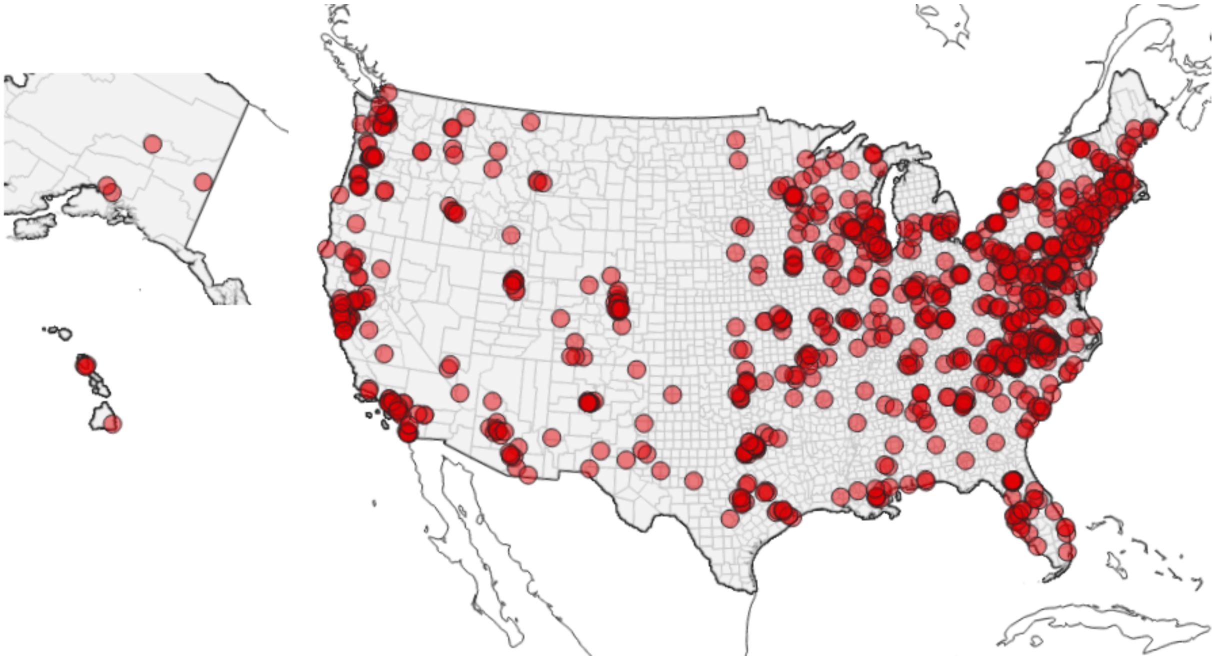

Figure E1: Locations of microbial samples. Each point represents the location of a home where samples of indoor and outdoor fungal communities was collected from the indoor and outdoor door sill. Samples were sequenced using ITS ribosomal RNA (rRNA) amplicon sequencing. All microbial data are publicly available and were downloaded on 21 July 2020. Additional details and references are given in the Methods (Supplementary Information).

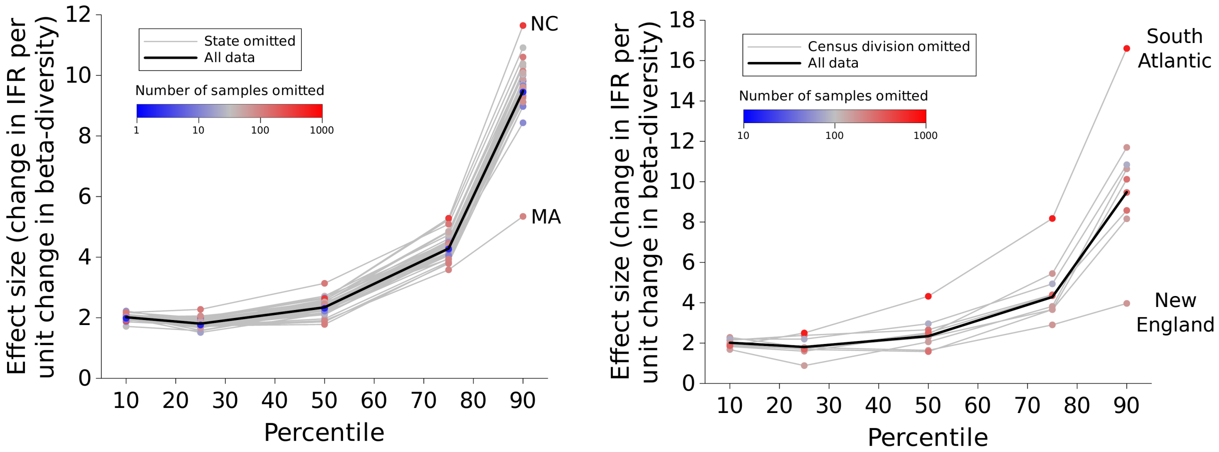


Figure E2: The associations observed between IFR and beta-diversity are not driven by a outlier states or United States Census Divisions. Removing the samples from each state (left) or United States Census Division (right) does not qualitatively change the association between IFR and beta-diversity (compare to the bold black line, which represents no omissions, and main text Figure 1A and 1B). The states and United States Census Divisions whose omissions have the greatest effects are noted. States and United States Census Divisions containing more microbial samples had larger effects on the results than those having fewer microbial samples.

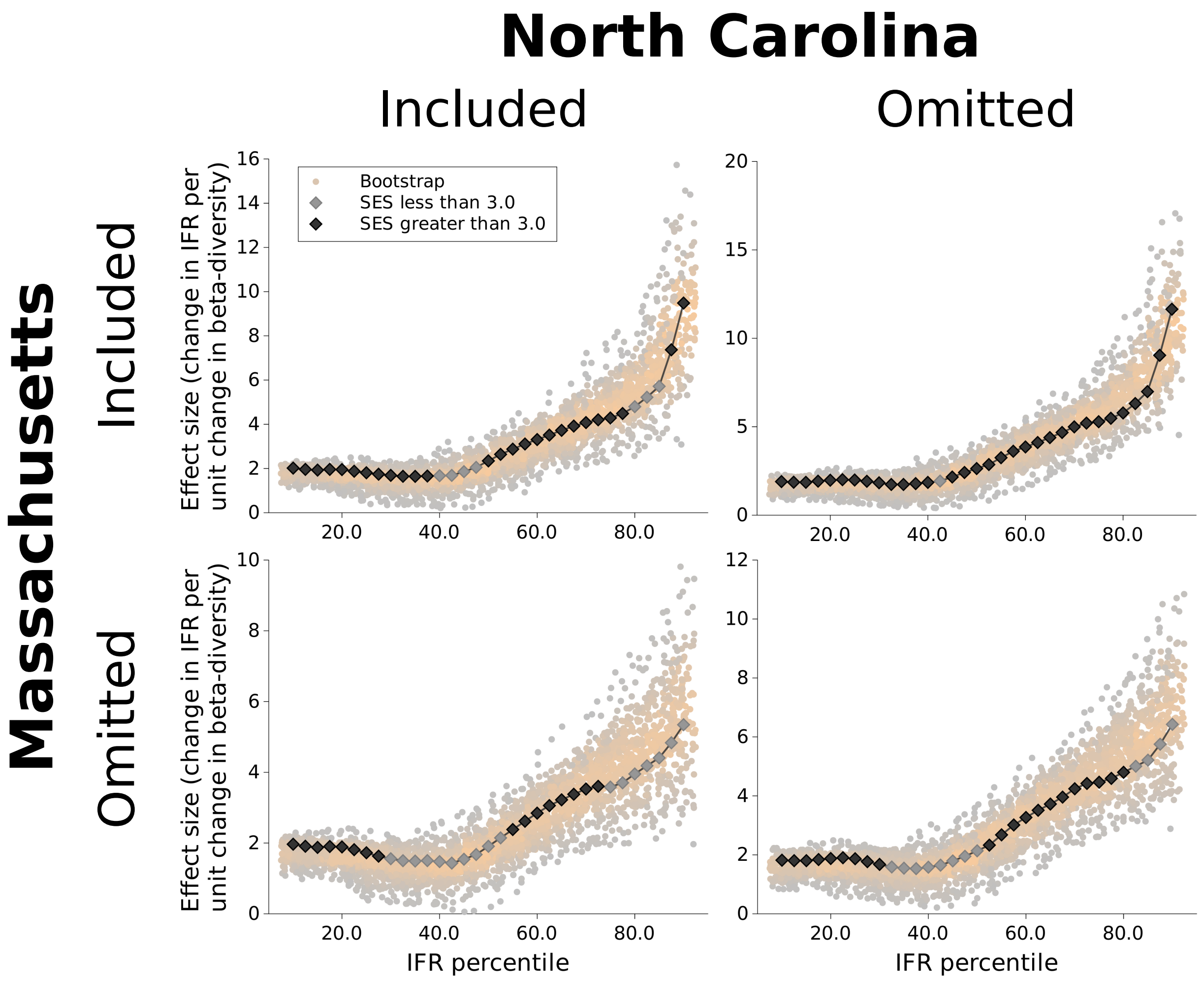


Figure E3: Effects of removing the data from Massachusetts and North Carolina on the association between IFR and beta-diversity. The data from Massachusetts and North Carolina have the greatest effects on the association between IFR and beta-diversity, in opposite directions (Figure E2). However, although it changes the magnitude of the effects, removing the data from (i) either of these states or (ii) both of these states does not change the overall nature of the association. Note the that scale of the $y$-axis differs between the graphs. The graph in the upper left, showing the associations for all of the data, is the same as the graph in main text Figure 1C, included again here for comparison.

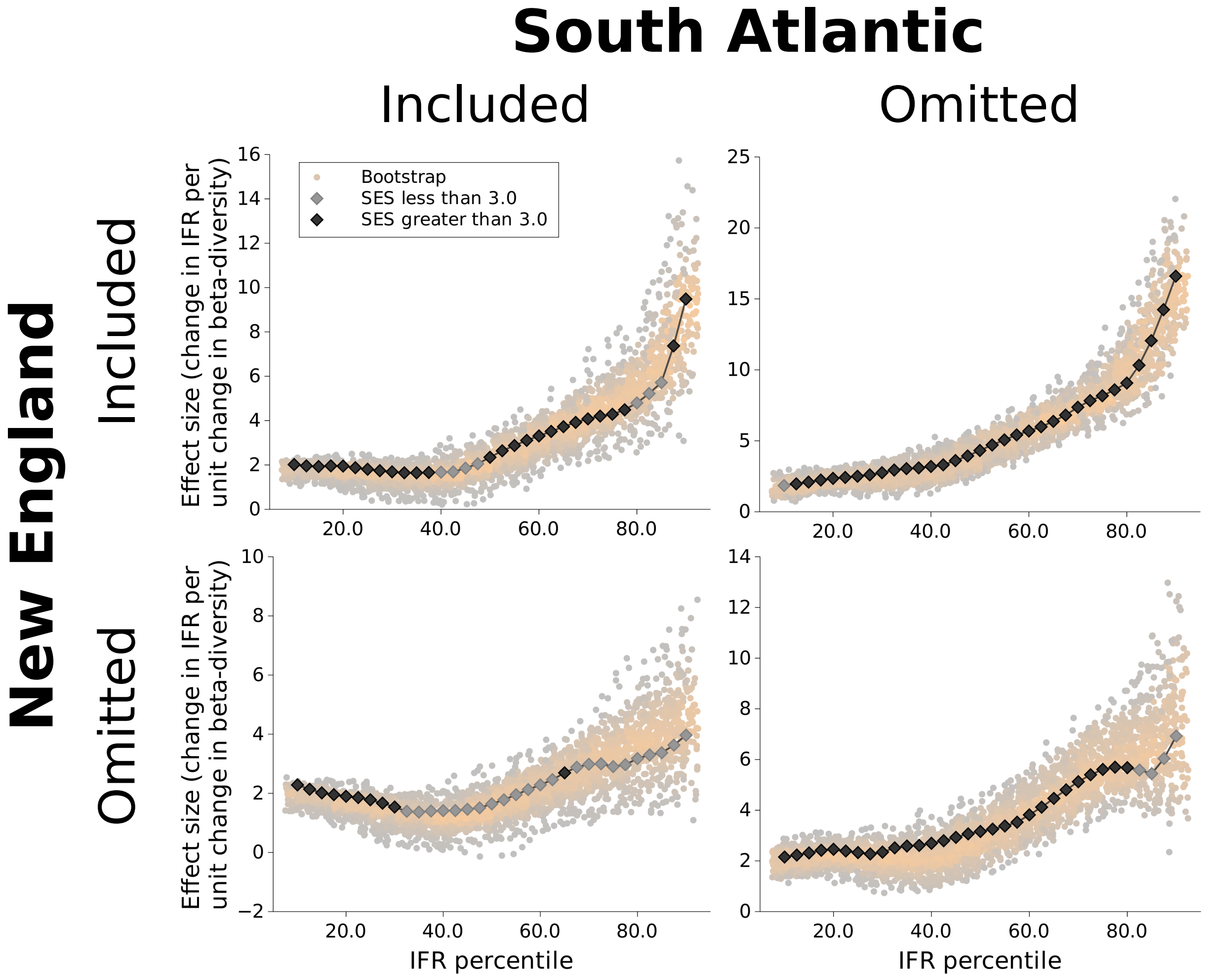


Figure E4: Effects of removing the data from New England and the South Atlantic on the association between IFR and beta-diversity. Of the data from the nine Census Regions in the United States, those from New England and the South Atlantic have the greatest effects on the association between IFR and beta-diversity (Figure E2). However, removing the data from the regions does not change the overall direction and trend of the association. Note the that scale of the $y$-axis differs between the graphs. The graph in the upper left, showing the associations for all of the data, is the same as the graph in main text Figure 1C, included again here for comparison.

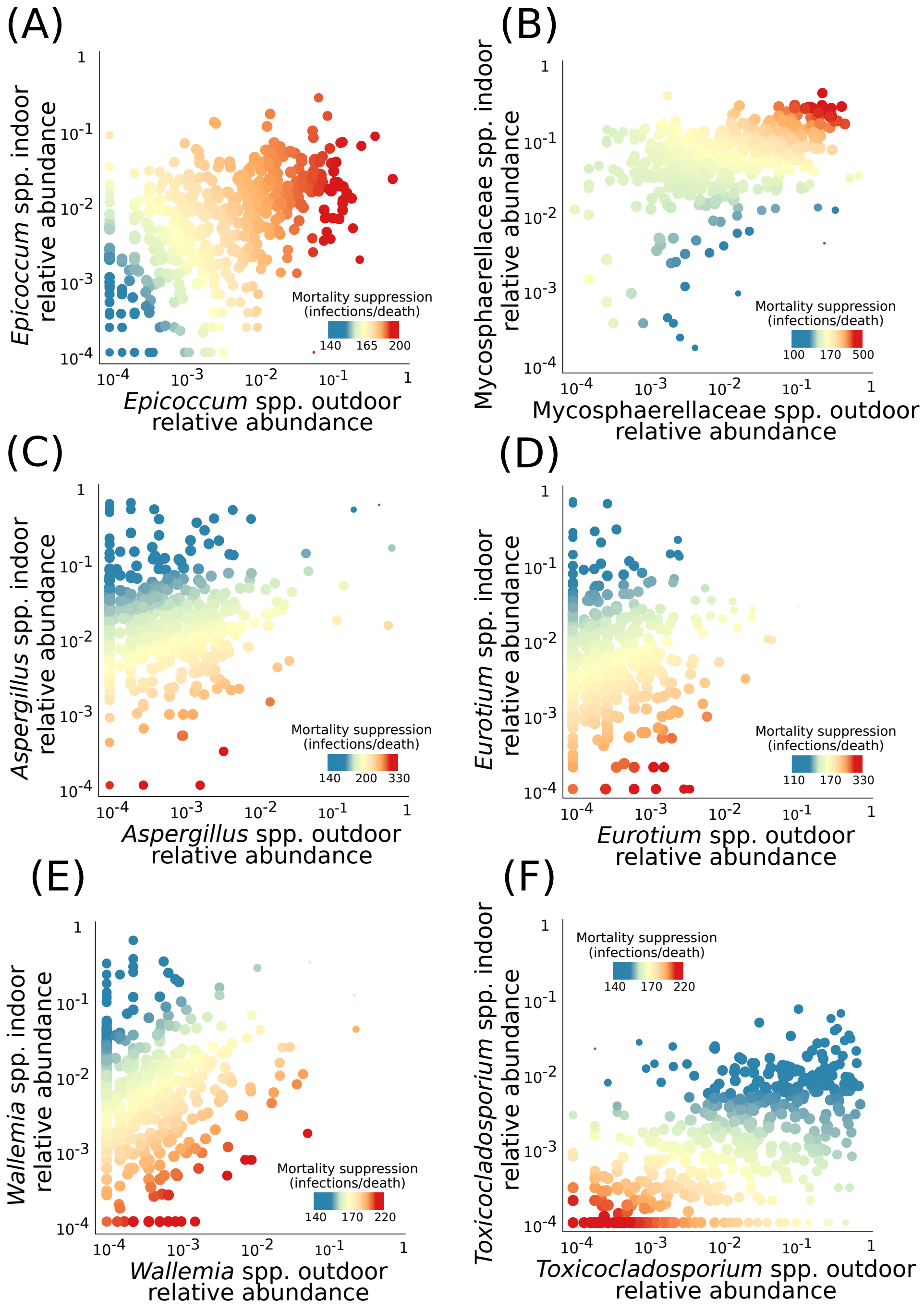


Figure E5: Association between indoor and outdoor relative abundance of selected genera and on COVID-19 mortality. COVID-19 mortality suppression is associated with high indoor and outdoor (A) *Epicoccum* and (B) Mycosphaerellaceae spp. relative abundance; relatively high outdoor but low indoor (C) *Aspergillus*, (D) *Eurotium*, and (E) *Wallemia* spp. relative abundance; and low indoor and outdoor (F) *Toxicocladosporium* spp. relative abundance. However, interactive effects dominate: collectively these genera suppress COVID-19 more than would be expected individually.

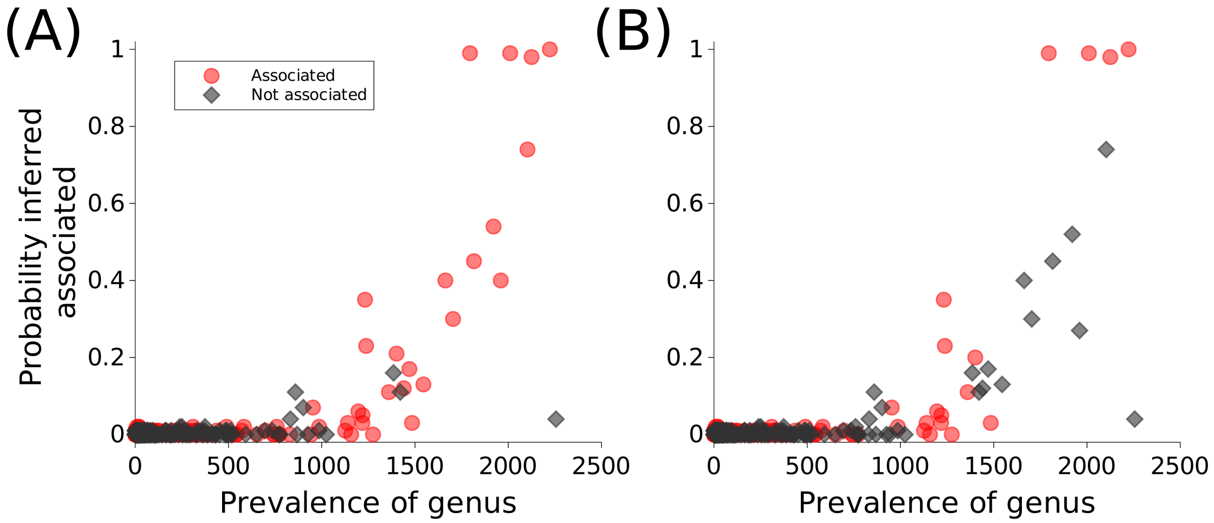


Figure E6: Performance of the method used to identify genera which drive the beta-diversity infection fatality rate association based on the analysis of simulated data where drivers are known. (A) The method correctly identifies most associated clusters with high prevalence genera and has low false positive rates. (B) A similar, but simpler method, wherein inferences are made about individual genera rather than clusters of genera, has much higher false positive rates.

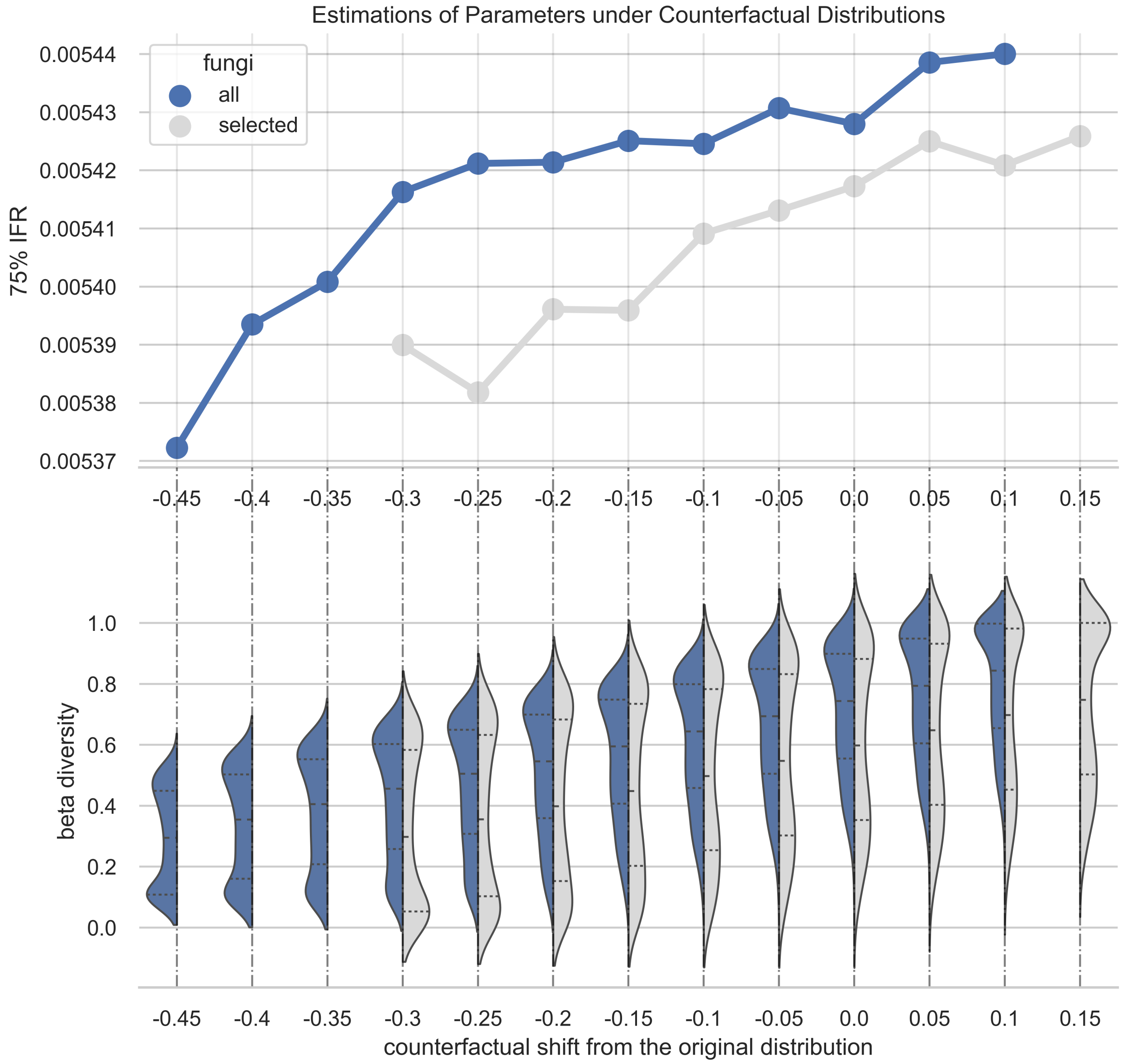


Figure E7: Point estimates of the 75-quantile IFR under different counterfactual distributions of fungi beta diversity, after accounting for other covariates.

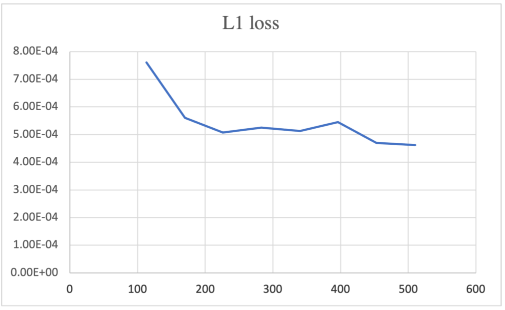


(a) L-1 loss under different sample size

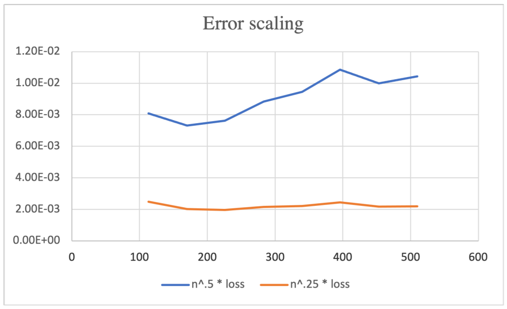


(b) loss scaling

Figure E8: Errors of the model for $E(Y \mid A, W)$ and its scaling when fitting with random forest. The result shows that the order of error is higher than $n^{-0.5}$ and the plug-in estimator will not be asymptotically normal in this case.
